# A Systematic Examination of Generative Artificial Intelligence (GAI) Usage Guidelines for Scholarly Publishing in Medical Journals

**DOI:** 10.1101/2024.03.19.24304550

**Authors:** Shuhui Yin, Peiyi Lu, Zhuoran Xu, Zi Lian, Chenfei Ye, Chihua Li

**Author notes:** Equally contributed as first authors (Shuhui Yin, Peiyi Lu, Zhuoran Xu). **Corresponding authors** Chihua Li, Institute for Social Research, University of Michigan, MI, USA 426 Thompson St, Ann Arbor, MI 48104; Chenfei Ye, International Research Institute for Artificial Intelligence, Harbin Institute of Technology (Shenzhen), Shenzhen, China.

## Abstract

**Background:** A thorough and in-depth examination of generative artificial intelligence (GAI) usage guidelines in medical journals will inform potential gaps and promote proper GAI usage in scholarly publishing. This study aims to examine the provision and specificity of GAI usage guidelines and their relationships with journal characteristics.

**Methods:** From the SCImago Journal Rank (SJR) list for medicine in 2022, we selected 98 journals as top journals to represent highly indexed journals and 144 as whole-spectrum sample journals to represent all medical journals. We examined their GAI usage guidelines for scholarly publishing between December 2023 and January 2024.

**Results:** Compared to whole-spectrum sample journals, the top journals were more likely to provide author guidelines (64.3% vs. 27.8%) and reviewer guidelines (11.2% vs. 0.0%) as well as refer to external guidelines (85.7% vs 74.3%). Probit models showed that SJR score or region was not associated with the provision of these guidelines among top journals. However, among whole-spectrum sample journals, SJR score was positively associated with the provision of author guidelines (0.85, 95% CI 0.49 to 1.25) and references to external guidelines (2.01, 95% CI 1.24 to 3.65). Liner models showed that SJR score was positively associated with the specificity level of author and reviewer guidelines among whole-spectrum sample journals (1.21, 95% CI 0.72 to 1.70), and no such pattern was observed among top journals.

**Conclusions:** The provision of GAI usage guidelines is limited across medical journals, especially for reviewer guidelines. The lack of specificity and consistency in existing guidelines highlights areas deserving improvement. These findings suggest that immediate attention is needed to guide GAI usage in scholarly publishing in medical journals.

**Key points:** *Question:* What are the provision and specificity of generative artificial intelligence (GAI) usage guidelines for scholarly publishing in top and whole-spectrum medical journals and their relationships with journal characteristics?

*Findings:* Author guidelines were more abundant and specific in top journals than in whole-spectrum journals. However, reviewer guidelines were extremely scarce in both groups of journals. Journal ranking score was associated with both provision and specificity of GAI usage guidelines in whole-spectrum journals while no significant relationship was found in top journals.

*Meaning:* The lack of provision and specificity as well as the inconsistencies in existing guidelines suggest that immediate attention is needed to guide GAI usage in scholarly publishing and safeguard integrity and trust in medical research.

## Introduction

ChatGPT, a chatbot powered by generative artificial intelligence (GAI) through large language models, was released in November 2022. It can generate responses based on statistical language patterns and is easily accessible to people without technical expertise.^1^ Its multifaceted capabilities and potential applications within diverse contexts have attracted billions of users and visits.^2–4^ Consequently, many researchers have been actively exploring ChatGPT and other similar tools’ potential applications to scholarly publishing, including manuscript preparation and peer review.^5–8^ To date, at least 1000 medical publications reported the use and impact of such tools in scholarly publishing.^9–11^ For manuscript preparation, researchers have used GAI tools to conduct literature review, formulate study design, analyze data, and even interpret results.^12–15^ For peer review, GAI has been used to summarize manuscript contents, review code, check methods, and even draft comments and feedback.^9,16^ These practices highlight that such tools have and will continue to transform scholarly publishing.

However, GAI is far from perfect and can lead to multiple concerns.^17–19^ Major concerns regarding its role in scholarly publishing include the eligibility of these tools for authorship, the risk of producing misleading or inaccurate information, breaches of data privacy and confidentiality, as well as challenges to integrity and originality.^20–23^ In response, many journals, organizations, and publishers have started to provide GAI usage guidelines for authors and reviewers and made continuous updates.^24–26^ For example, medical publication organizations, and publishers, such as the International Committee of Medical Journal Editors (ICMJE) and Elsevier, have decided that GAI should not be listed as authors because they cannot be accountable for the submitted work;^27,28^ some journals require authors to document details of GAI usage and fact-check GAI-generated contents; and some journals strictly prohibit the use of GAI in peer review.^29^ These developments show the new challenges posed by GAI and the scientific community’s corresponding adaptations.

Despite a fast-growing increase in discussions on GAI usage in scientific research, few have examined GAI usage guidelines systematically. For example, one study examined GAI usage guidelines for authors based on the 100 largest publishers and top 100 highly ranked journals of different disciplines, and it found less than a quarter of these publishers and around 85% of the top journals provided author guidelines.^30^ The other study examined author disclosure requirements for GAI usage in 125 nursing journals and found less than 40% of them had explicit instructions.^31^ They offered valuable insights into the evolving ethical standards and practices of this rapidly changing field.

However, many other important aspects of GAI usage guidelines remain less explored or unknown. First, few or no existing studies examined GAI guidelines for reviewers, ignoring the equal importance of peer review in scholarly publishing. Second, previous studies either focused on top-ranked journals or only on nursing journals, leading to an incomplete assessment of such guidelines across medical journals. Third, little exploration has been made on the relationship between journal characteristics and GAI guidelines for authors and reviewers. To address these gaps, our study focuses on medical journals and includes both top and whole-spectrum journals. We aim to delineate the provision and specificity of GAI guidelines for authors and reviewers as well as external guidelines referred by these journals. We will also examine potential relationships between journal characteristics and the provision and specificity of different GAI guidelines. These together will provide a thorough overview of current practices and contribute to the ongoing development of GAI usage guidelines across medical journals.

## Methods

### Journal selection

Based on the SCImago Journal Rank (SJR), two lists of journals were selected from the journal ranking list for medicine in 2022: ‘top’ journals and whole-spectrum sample journals (**Figure 1**).^32^ Each journal’s ranking was determined by its corresponding SJR score, a measure of the number of citations received by its articles that contextualizes the journal’s prestige and popularity within the academic community.

**Figure 1.**
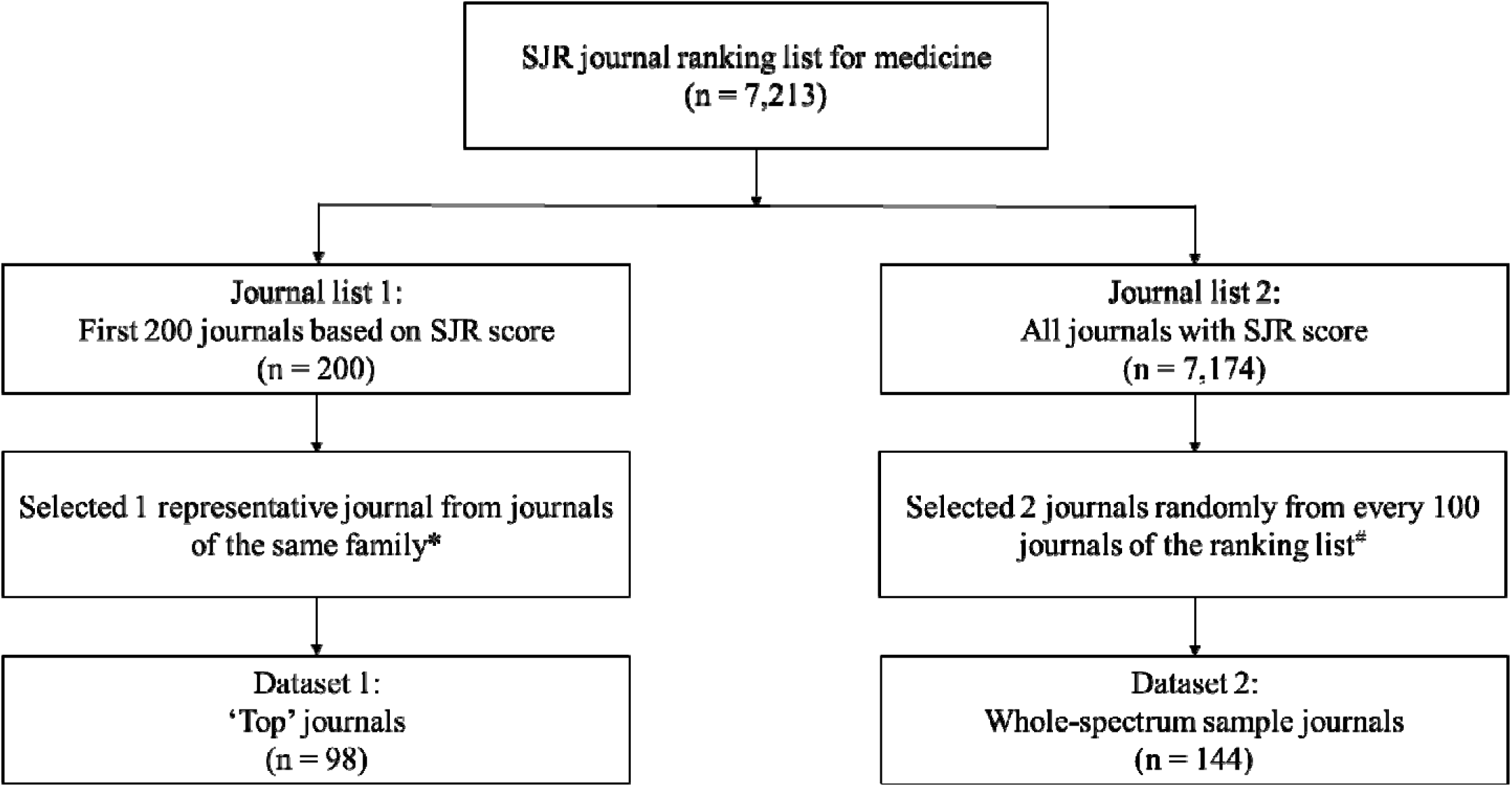
Flowchart of journal selection. Footnote: * Journals from the same family most likely adopted identical guidelines for GAI usage. # Non-English journals were excluded; Journals from the same family were replaced.

The list of top journals was generated due to the observation that many leading journals had extensive discussions on how to use GAI or relevant tools in scholarly publishing, the inclusion of which may provide rich information on such guidelines.^7,33^ To obtain a list of top journals, we selected a representative journal with the highest SJR score from each journal family among the first 200 ranked journals. This is because journals within the same family usually adopt identical GAI usage guidelines. For example, the Lancet and other 21 Lancet-affiliated journals were all among the first 200 ranked journals and adopted identical guidelines.

The list of whole-spectrum sample journals was generated to comprehensively examine the current state of GAI usage guidelines across medical journals. For this journal list, we randomly selected two journals from every 100 journals on the ranking list. Non-English journals were excluded, and journals of the same family were replaced. As a result, we included 98 journals for top journals and 144 journals for whole-spectrum sample journals.

### Data collection and extraction

For each journal selected above, two members of our research team independently carried out a thorough examination of the guidelines, which were publicly available on each journal’s website. This was to identify any content related to the GAI usage. The following information was extracted to create two datasets for top journals and whole-spectrum sample journals respectively: journal characteristics and three types of GAI usage guidelines (**Table 1**). Journal characteristics included three items: journal name, SJR score, and region. Three types of GAI usage guidelines included author guidelines, reviewer guidelines, and references to external guidelines. All the data extraction was completed between December 2023 and January 2024. Any disagreements were resolved through group discussions.

**Table 1.** Journal characteristics and three types of generative artificial intelligence (GAI) usage guidelines extracted.

| Items | Journal characteristics | Three types of GAI usage guidelines |  |  |
| --- | --- | --- | --- | --- |
|  |  | Author guidelines (Y/N/NR) | Reviewer guidelines (Y/N/NR) | References to external guidelines (Y/NR) |
| 1 | Journal name | Usage permission | Usage permission | COPE |
| 2 | SJR score | Language editing | Language editing | ICMJE |
| 3 | Region | Manuscript writing | Usage documentation | WAME |
| 4 |  | Data analysis and interpretation |  | Publishers |
| 5 |  | Image generating |  |  |
| 6 |  | Fact-checking requirement |  |  |
| 7 |  | Usage documentation |  |  |
| 8 |  | Authorship eligibility |  |  |

Footnote: Abbreviations: Y, Yes; N, No; NR, Not Reported;
COPE, Committee on Publication Ethics; ICMJE, International Committee of Medical Journal Editors; WAME, World Association of Medical Editors.

For author guidelines, we identified eight requirements: usage permission, language editing, manuscript writing, data analysis and interpretation, image generating, fact-checking requirement, usage documentation, and authorship eligibility. Usage permission refers to whether GAI tools are allowed to be used in manuscript preparation. Language editing, manuscript writing, data analysis and interpretation, and image generating refer to whether GAI tools are allowed to be used in each of these manuscript development steps. Fact-checking requirement refers to whether authors are required to check and verify AI-generated content. Usage documentation refers to whether authors are required to disclose and document the use of GAI tools. Authorship eligibility refers to whether AI owns authorship and is allowed to be listed as an author. Journals providing any of these eight requirements were marked as providing author guidelines.

For reviewer guidelines, we identified three requirements: usage permission, language editing, and usage documentation. Journals providing any of these three requirements were marked as providing reviewer guidelines. We further coded each of the 11 requirements for author and reviewer guidelines as ‘Yes’, ‘No’, or ‘Not Reported’.

For each journal, we generated a specificity score for author guidelines (range: 0 to 8), reviewer guidelines (range: 0 to 3), and author and reviewer guidelines combined (range: 0 to 11), respectively. Those scores were derived from the above coding: a score of ‘1’ was assigned for either ‘Yes’ or ‘No’ to each requirement; and a score of ‘0’ for “Not Reported”. A higher specificity score indicated a higher specificity level.

Many journals referred to external guidelines formulated by publishing organizations or publishers for authors and reviewers. For these external guidelines on GAI usage, we focused on standards formulated by four groups: the Committee on Publication Ethics (COPE), ICMJE, the World Association of Medical Editors (WAME), and publishers. We coded the reference to each of these four external guidelines as ‘Yes’ or ‘Not Reported’. We did not examine the specificity score of external guidelines because they were not journal-specific.

### Statistical analysis

We summarized and compared the characteristics and provision of different usage guidelines between top journals and whole-spectrum sample journals. We used Probit regression to examine relationships between journal characteristics and the provision of author or reviewer guidelines and references to external guidelines. We used linear regression to relationships between journal characteristics and the specificity score of these guidelines. We conducted all analyses using R 4.2. All data used for this study was provided in **Supplementary Tables 1 and 2**.

## Results

**Table 2** summarizes journal characteristics and the provision of different GAI usage guidelines. The 98 top journals had a median SJR score of 4.2, and 95 of them (96.9%) were based in either Northern America or Western Europe. The 144 whole-spectrum sample journals had a median SJR score of 0.5, and they were more evenly distributed across the three regions. The top journals were more likely to provide different types of GAI usage guidelines than whole-spectrum sample journals, including author guidelines (64.3% vs. 27.8%), reviewer guidelines (11.2% vs. 0.0%), and references to external guidelines (85.7% vs 74.3%). A detailed summary of the provision and specificity of GAI usage guidelines is presented for top journals and whole-spectrum sample journals in **Supplementary Table 3**, including eight requirements for authors, three requirements for reviewers, and four frequently referred external guidelines. Compared to whole-spectrum sample journals, top journals had a higher specificity level of author guidelines, reviewer guidelines, and the two combined.

**Table 2.** Journal characteristics and provision of different GAI usage guidelines.

|  | Top journals<br>(n = 98) | Whole-spectrum sample<br>journals (n = 144) | P value<sup>#</sup> |
| --- | --- | --- | --- |
| Journal characteristics |  |  |  |
| SJR score, median (IQR) | 4.2 (3.6, 6.7) | 0.5 (0.2, 0.9) | < 0.01 |
| Region, n (%) |  |  |  |
| Northern America | 50 (51.0) | 39 (27.1) | < 0.01 |
| Western Europe | 45 (45.9) | 51 (35.4) |  |
| Other regions* | 3 (3.1) | 54 (37.5) |  |
| Provision of different usage guidelines |  |  |  |
| Author guidelines, n (%) | 63 (64.3) | 40 (27.8) | < 0.01 |
| Reviewer guidelines, n (%) | 11 (11.2) | 0 (0.0) | < 0.01 |
| References to external guidelines, n (%) | 84 (85.7) | 107 (74.3) | < 0.05 |

Footnote: \*Other regions included Africa, Asiatic region, Eastern Europe, Latin America, Middle East, and Pacific region.
<sup>#</sup> Differences between the two groups of journals were analyzed using Wilcoxon Rank Sum test or Chi-Square tests.

Relationships between journal characteristics and the provision of GAI usage guidelines among top journals differed from those among whole-spectrum sample journals (**Table 3**). Among top journals, the provision of these guidelines generally showed no significant differences comparing journals with varying SJR scores or those based in different regions. However, journals based in Western Europe were more likely to refer to external guidelines than those based in Northern America (2.07, 95% CI: 1.07 to 4.26). Among whole-spectrum sample journals, journals with a higher SJR score were more likely to provide author guidelines (0.85, 95% CI: 0.49 to 1.25) and refer to external guidelines (2.01, 95% CI: 1.24 to 3.65). Compared to journals based in Northern America, those in Western Europe had a similar likelihood of providing author guidelines (0.02, 95% CI: −0.51 to 0.55), whereas journals in other regions were less likely to do so (−1.03, 95% CI: −1.67 to −0.43). Since no reviewer guidelines were provided by whole-spectrum sample journals, no association estimates were obtained.

**Table 3.**
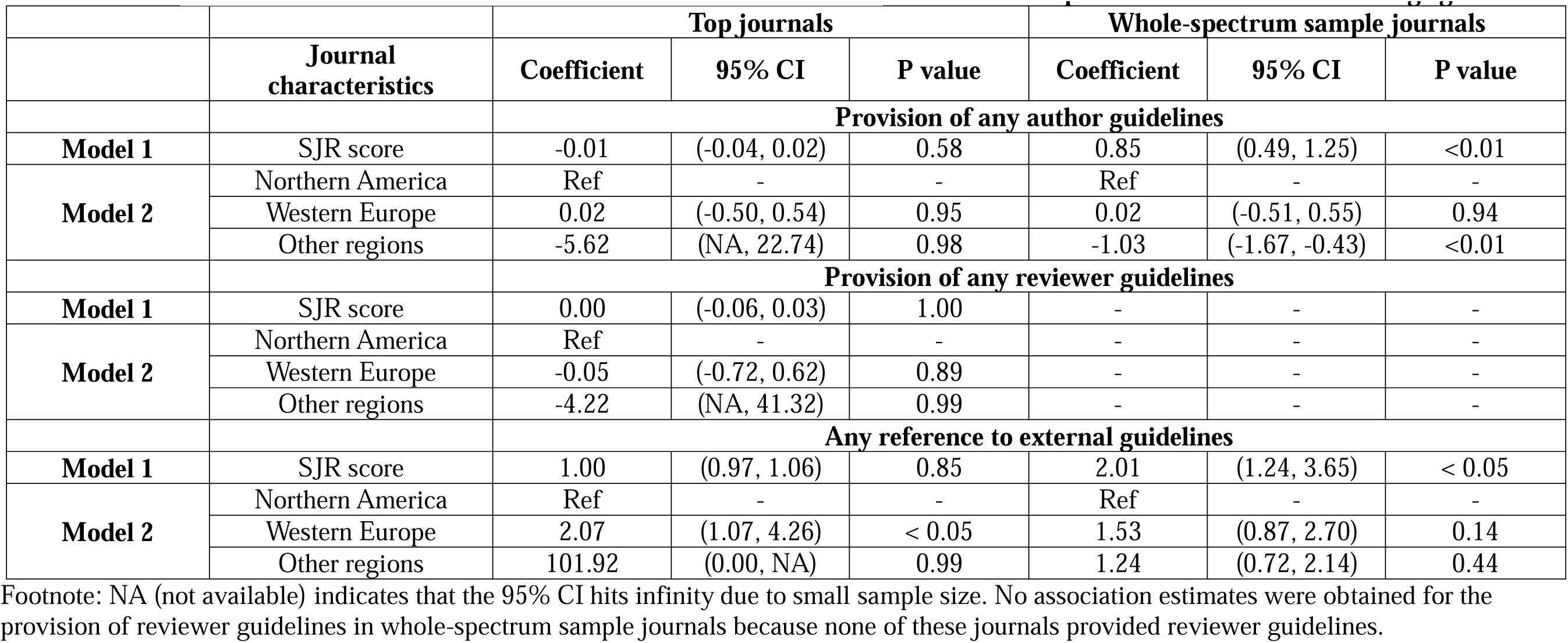
Probit regression analysis of the relationship between journal characteristics and the provision of different GAI usage guidelines.

|  |  | Top journals |  |  | Whole-spectrum sample journals |  |  |
| --- | --- | --- | --- | --- | --- | --- | --- |
|  | Journal characteristics | Coefficient | 95% CI | P value | Coefficient | 95% CI | P value |
|  |  | **Provision of any author guidelines** |  |  |  |  |  |
| **Model 1** | SJR score | -0.01 | (-0.04, 0.02) | 0.58 | 0.85 | (0.49, 1.25) | <0.01 |
| **Model 2** | Northern America | Ref | - | - | Ref | - | - |
|  | Western Europe | 0.02 | (-0.50, 0.54) | 0.95 | 0.02 | (-0.51, 0.55) | 0.94 |
|  | Other regions | -5.62 | (NA, 22.74) | 0.98 | -1.03 | (-1.67, -0.43) | <0.01 |
|  |  | **Provision of any reviewer guidelines** |  |  |  |  |  |
| **Model 1** | SJR score | 0.00 | (-0.06, 0.03) | 1.00 | - | - | - |
| **Model 2** | Northern America | Ref | - | - | - | - | - |
|  | Western Europe | -0.05 | (-0.72, 0.62) | 0.89 | - | - | - |
|  | Other regions | -4.22 | (NA, 41.32) | 0.99 | - | - | - |
|  |  | **Any reference to external guidelines** |  |  |  |  |  |
| **Model 1** | SJR score | 1.00 | (0.97, 1.06) | 0.85 | 2.01 | (1.24, 3.65) | < 0.05 |
| **Model 2** | Northern America | Ref | - | - | Ref | - | - |
|  | Western Europe | 2.07 | (1.07, 4.26) | < 0.05 | 1.53 | (0.87, 2.70) | 0.14 |
|  | Other regions | 101.92 | (0.00, NA) | 0.99 | 1.24 | (0.72, 2.14) | 0.44 |

Footnote: NA (not available) indicates that the 95% CI hits infinity due to small sample size. No association estimates were obtained for the provision of reviewer guidelines in whole-spectrum sample journals because none of these journals provided reviewer guidelines.

Figure 2 shows the specificity level of author and reviewer guidelines and detailed requirements of different guidelines among top journals. The specificity score was determined by how many of the 11 requirements of author and reviewer guidelines were provided. For author guidelines, a consensus emerged on five requirements whenever provided: permitting GAI usage, allowing for language editing, requiring fact-checking, mandating usage documentation, and prohibiting AI authorship. However, disagreements were observed about whether GAI can be used for writing manuscripts, analyzing and interpreting data, and generating images. For reviewer guidelines, fewer journals provided requirements, and there were disagreements on permitting GAI usage and allowing for language editing. Among external guidelines, the ICMJE guideline was the most frequently referred to. Details of external guidelines are summarized in **Supplementary Table 4**. Among whole-spectrum sample journals, similar patterns of requirements were observed except that a lower proportion of journals provided specific requirements (**Supplementary Figure 1**).

**Figure 2.**
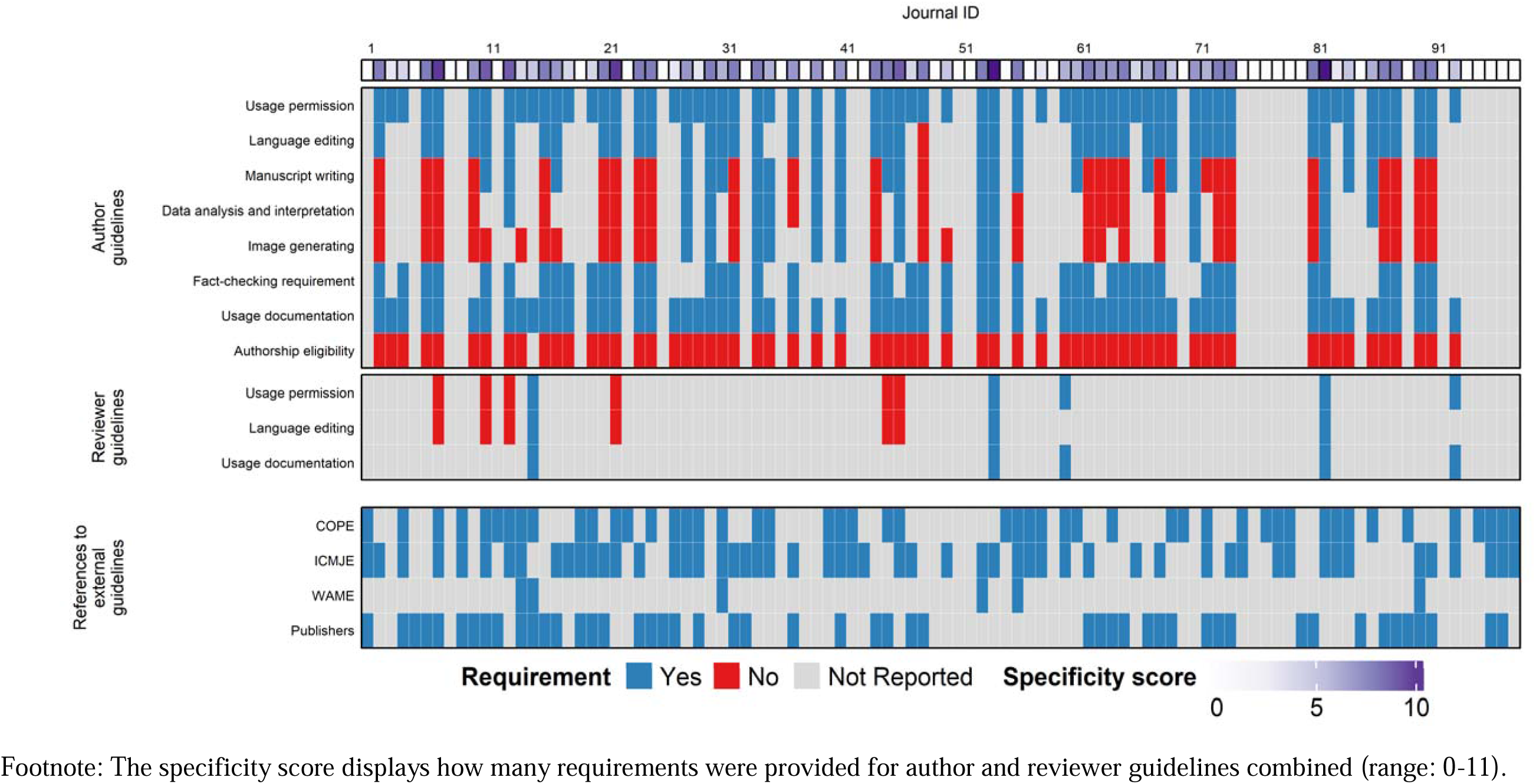
Specificity level of author and reviewer guidelines and requirements of different GAI usage guidelines among top journals. Footnote: The specificity score displays how many requirements were provided for author and reviewer guidelines combined (range: 0-11).

Relationships between journal characteristics and specificity scores of author and reviewer guidelines were examined (**Supplementary Table 5**). Among top journals, the journal’s SJR score was not associated with the specificity level of usage guidelines. In contrast, whole-spectrum sample journals with a higher SJR score exhibited a higher specificity level. Among both groups of journals, those based in Western Europe had a similar level of specificity as those in North America, whereas journals from other regions had a lower specificity level. Relationships for author guidelines showed similar patterns among top journals.

## Discussion

Our study systematically examined GAI usage guidelines for scholarly publishing across medical journals, including author guidelines, reviewer guidelines, and references to external guidelines. Around two-thirds of top journals provided author guidelines while less than one-third of whole-spectrum journals did so, highlighting differential acknowledgment and integration of GAI practices and room for improvement across medical journals. The extremely low proportion of both top and whole-spectrum sample journals providing reviewer guidelines showed their negligence in setting guidance on the use of such tools for reviewers. The high proportion of both journal groups referred to external guidelines showed the critical role of these external standards in fostering ethical practices of GAI usage. We also identified journal characteristics that were associated with the provision and specificity level of different guidelines. Together, our results suggest the urgent need for explorations and improvements of GAI usage guidelines in scholarly publishing.

Among medical journals across the whole spectrum, only a small proportion of journals (28%) provided GAI usage guidelines for authors, highlighting a serious lack of alertness in guiding the proper use of such tools. Lack of proper guidance can lead to unethical practices or allow AI-generated manuscripts to infiltrate publications undetected, jeopardizing the quality, integrity, and transparency of research.^34–36^ Interestingly, the figure we observed is considerably lower than that reported by another study (86%) focusing on only top journals.^30^ Moreover, the provision of author guidelines among top medical journals (64%) is also lower in our study compared to that study. The main reason for the discrepancy in the provision of author guidelines between our study and the other is the different journal selection methods. Our study selected one representative journal from each journal family while the other study included multiple journals of the same families. For example, among their 51 top medical journals, six were Nature family journals and three were JAMA family journals, which usually adopted the same guidelines. This journal selection method can lead to biases in examining the provision of GAI usage guidelines. Indeed, we observed a similar proportion of journals providing author guidelines among the first 200 SJR-ranked medical journals when we included multiple journals of the same families.

The better provisions of author guidelines comparing top journals to whole-spectrum journals can be attributed to several factors. They include more accessible advanced AI technology in Western Europe and Northern America, stringent regulatory environments, and robust scholarly communication networks.^37–39^ Author guidelines provided by top journals are also more specific than others, evidenced by two observations. First, top journals had a much higher specificity level than whole-spectrum sample journals; second, among whole-spectrum sample journals, journals with a higher SJR score tend to be more specific. This greater specificity level can help authors adhere to ethical standards and prevent potential misconduct, including plagiarism or misrepresentation of AI-generated content as human work. Our findings showed that both top and whole-spectrum journals have significant room for improvement.

Across journals, consensus has been achieved for five of the eight requirements examined in author guidelines, and disagreements were observed for three requirements related to content generating applications. The remarkable capacities of GAI justify its usage in scholarly writing, especially for language editing.^40–43^ However, its risks threatening research integrity require authors to conduct fact-checking and usage documentation but not to grant such tools authorship.^44^ These agreements show how the scientific community is embracing GAI in a cautious way. For example, the emphasis on fact-checking and usage documentation is necessary for preventing fabrication and potential biases and promoting transparency and trust.^45^

Notably, journals differ in instructions for usage documentation on what should be disclosed. While some journals require authors to report any use of GAI, other journals allow authors not to do so if such tools were solely used for language editing. This may be explained by journals’ different interpretations of GAI applications. Together with two other recent studies showing inconsistencies in what to disclose of GAI usage, a need is highlighted surrounding standardization of GAI usage disclosure and documentation.^30,31,46^ More importantly, journals held different and even opposite stances for content-generating applications, including manuscript writing, data analysis and interpretation, and image generating. This is due to concern centered on plagiarism because it is challenging to trace and verify the originality of AI-generated content.^47,48^

As the first study that systematically examined GAI usage guidelines for reviewers, we are surprised to find that an extremely small proportion of journals did so. For example, only 10% of top journals and none of the whole-spectrum sample journals provided reviewer guidelines. Given the reviewers’ pivotal role in safeguarding the quality and integrity of scholarly work, this negligence reflects a concerning gap in guiding the use of such tools in the peer review process. The misuse of GAI in peer review can have multiple negative consequences, including the breach of private data and violation of intellectual property. These are important issues considering the sensitive nature of health data and medical research.^22^

Among the merely available reviewer guidelines identified, we observed disagreements on requirements regarding usage permission, language editing, and usage documentation across journals. We interpreted these disagreements as journals’ different stances on how to protect the confidentiality of manuscripts.^49,50^ Some journals specified conditions in which GAI can be used. For example, they required reviewers to obtain permission from authors and editors, to confirm that manuscripts shall not be used as training data, and provide authors with the choice to opt for or against a GAI-assisted review process.^26,51^

Around 90% of top journals and three quarters of whole-spectrum sample journals have referred to external guidelines on GAI usage formulated by ICMJE, COPE, WAWE, or publishers. This underscores a collective endeavor to establish ethical benchmarks for GAI usage in scholarly publishing. Notably, top journals based in Western Europe were more likely to refer to external standards than their Northern American counterparts. This may stem from Western Europe’s pioneering regulations on GAI usage.^52,53^ Importantly, we identified conflicts between journal-specific guidelines and referred external guidelines for at least five of the top journals we selected. For example, *Molecular Aspects of Medicine* prohibits content-generating applications, while its referred external guideline, WAME, permits such usage. These conflicts have also been reported by another study.^30^ To avoid confusion, we call for attention to resolve such conflicts.

We identified three main aspects of GAI usage guidelines that need urgent improvements. First, it is important for journals to provide specific GAI usage guidelines for both authors and reviewers rather than only refer to external guidelines. Journal-specific guidelines can better ensure the relevance and effectiveness in guiding GAI usage in their unique scholar environment. Second, details of the guidelines should be specified to avoid confusion, especially for content-generating applications. Our systematic evaluation of available GAI usage guidelines provides a roadmap to clarify expectations on proper GAI practices. Third, disagreements identified across guidelines warrant special attention. Although these disagreements highlight the continuously evolving use of GAI, it is possible to establish a framework of proper practices and specify requirements under different scenarios. For example, journals may consider standardizing usage documentation by integrating questions into the manuscript submission system; ascertaining whether GAI was used, and if so, detailing specific purposes and ensuring accountability. Besides, it is essential to regularly review, discuss, and update GAI usage guidelines.

Our study has several strengths. It pioneered a systematic and quantitative examination of GAI usage guidelines across top and whole-spectrum medical journals, which provides an overview of the current regulations in scholarly publishing. It also synthesized existing GAI usage guidelines for both authors and reviewers and established a systematic checklist. We further provided recommendations aimed at improving existing GAI usage guidelines for scholarly publishing and promoting the proper use of GAI tools. While our study focused on medical journals, we believe our findings and recommendations can be applied to journals in other disciplines.

Our study also has limitations. First, for whole-spectrum sample journals, we sampled 2% of the 7,174 medical journals with an SJR score and included only English journals. Despite this, our use of stratified random sampling enables the representation of all medical journals listed in the SJR database and allows for timely communication of our important findings. Second, the scarce provision of reviewer guidelines observed in our findings might be attributed to the potential non-public accessibility of reviewer guidelines, which we think is uncommon. Third, our analysis did not include guidelines for editors, despite their relevance and importance, due to their scarce availability across the journals we examined. In the future, we would like to collaborate with different journals to conduct such examinations.

## Conclusions

The provision of GAI usage guidelines is limited across medical journals, especially for reviewer guidelines. The lack of specificity and inconsistencies in existing guidelines highlight areas deserving improvement. Immediate attention is needed to guide GAI usage in scholarly publishing and safeguard the integrity and trust of medical research.

## Data Availability

All data relevant to the study are included in the article or included as supplementary materials. Full data and analytical code are available from the corresponding author at.

## Corresponding Author

Chenfei Ye, Chihua Li.

## Author Contributions

Li had full access to all of the data in the study and took responsibility for the integrity of the data and the accuracy of the data analysis.

*Concept and design:* Yin, Li.

*Acquisition, analysis, or interpretation of data:* All authors.

*Drafting of the manuscript:* Yin, Li.

*Critical review of the manuscript for important intellectual content:* All authors.

*Statistical analysis:* All authors.

*Obtained funding:* Ye, Li.

*Administrative, technical, or material support:* Yin, Li.

*Supervision:* Li.

## Conflict of Interest Disclosures

None.

## Funding/Support

Chenfei Ye is supported by grants from the National Natural Science Foundation of P.R. China (62106113); Guangdong Basic and Applied Basic Research Foundation (2023A1515010792), Basic Research Foundation of Shenzhen Science and Technology Stable Support Program (GXWD 20231129121139001).

## Role of the Funder/Sponsor

The funders had no role in the design and conduct of the study; collection, management, analysis, and interpretation of the data; preparation, review, or approval of the manuscript; and decision to submit the manuscript for publication.

## Notes

### Competing Interest Statement

The authors have declared no competing interest.

