## Supplementary Tables and Figures for "A Systematic Examination of Generative Artificial Intelligence (GAI) Usage Guidelines for Scholarly Publishing in Medical Journals"

### Supplementary Material

**Table of Contents**

**Supplementary Table 1. Journal characteristics and information sources of generative artificial intelligence (GAI) usage guidelines of top journals**

**Supplementary Table 2. Journal characteristics and information sources of GAI usage guidelines of whole-spectrum sample journals**

**Supplementary Table 3. Provision of specific requirements of different GAI usage guidelines**

**Supplementary Table 4. External GAI usage guidelines and their requirements**

**Supplementary Table 5. Linear regression analysis of the relationship between journal characteristics and the specificity score of GAI usage guidelines**

**Supplementary Figure 1. Specificity level of author and reviewer guidelines and requirements of different GAI usage guidelines among whole-spectrum sample journals**

**Supplementary Table 1. Journal characteristics and information sources of GAI usage guidelines of top journals**

| ID | Journal name | SJR Score | Region | Website |
|---|---|---|---|---|
| 1 | Ca-A Cancer Journal for Clinicians | 86.09 | Northern America | https://acsjournals.onlinelibrary.wiley.com/hub/journal/15424863/homepage/forauthors.html; https://authorservices.wiley.com/Reviewers/journal-reviewers/index.html |
| 2 | New England Journal of Medicine | 26.02 | Northern America | https://www.nejm.org/about-nejm/editorial-policies |
| 3 | Nature Medicine | 24.69 | Western Europe | https://www.nature.com/nm/submission-guidelines |
| 4 | MMWR Recommendations and Reports | 23.96 | Northern America | https://www.cdc.gov/mmwr/author_guide.html |
| 5 | Annual Review of Immunology | 15.26 | Northern America | https://www.annualreviews.org/page/authors/editorial-policies#authorship |
| 6 | Immunity | 14.80 | Northern America | https://www.cell.com/immunity/authors |
| 7 | Lancet. The | 14.61 | Western Europe | https://www.thelancet.com/lancet/about; https://www.thelancet.com/peer-review |
| 8 | World Psychiatry | 14.31 | Northern America | https://onlinelibrary.wiley.com/page/journal/20515545/homepage/forauthors.html |
| 9 | Physiological Reviews | 13.69 | Northern America | https://journals.physiology.org/ethics |
| 10 | Annals of Oncology | 11.95 | Western Europe | https://www.sciencedirect.com/journal/annals-of-oncology/publish/guide-for-authors |
| 11 | Science Immunology | 11.19 | Northern America | https://www.science.org/content/page/science-immunology-information-authors#editorial-policies; https://www.science.org/content/page/science-journals-editorial-policies#general-policies |
| 12 | Current Protocols in Bioinformatics | 10.41 | Northern America | https://currentprotocols.onlinelibrary.wiley.com/hub/authorguidelines |
| 13 | Journal of Clinical Oncology | 10.16 | Northern America | https://ascopubs.org/jco/authors/journal-policies; https://ascopubs.org/jco/authors/journal-policies#AI |
| 14 | Molecular Cancer | 8.70 | Western Europe | https://www.biomedcentral.com/getpublished/editorial-policies#artificial+intelligence+%28ai%29 |
| 15 | Gut | 8.59 | Western Europe | https://gut.bmj.com/pages/authors#submission_guidelines; https://authors.bmj.com/policies/ai-use/ |
| 16 | Journal of the American College of Cardiology | 8.34 | Northern America | https://www.sciencedirect.com/journal/journal-of-the-american-college-of-cardiology/publish/guide-for-authors |
| 17 | JAMA Oncology | 8.10 | Northern America | https://jamanetwork.com/journals/jamaoncology/pages/instructions-for-authors |
| 18 | Circulation | 7.80 | Northern America | https://www.ahajournals.org/submission-requirements |
| 19 | Gastroenterology | 7.65 | Western Europe | https://www.gastrojournal.org/content/authorinfo |
| 20 | Clinical Microbiology Reviews | 7.58 | Northern America | https://journals.asm.org/authorship |
| 21 | Journal of Hepatology | 7.40 | Western Europe | https://www.journal-of-hepatology.eu/content/authorinfo |
| 22 | Cancer Discovery | 7.27 | Northern America | https://aacrjournals.org/pages/editorial-policies; https://aacrjournals.org/pages/editorial-process#peerrev |
| 23 | Endocrine Reviews | 7.21 | Northern America | https://academic.oup.com/edrv/pages/author_guidelines |
| 24 | European Urology | 6.96 | Western Europe | https://www.sciencedirect.com/journal/european-urology/publish/guide-for-authors |
| 25 | Chem | 6.80 | Northern America | https://www.cell.com/chem/authors; https://www.elsevier.com/about/policies-and-standards/publishing-ethics#Authors |
| 26 | Accounts of Chemical Research | 6.38 | Northern America | https://pubs.acs.org/page/policy/ethics/index.html |
| 27 | American Journal of Respiratory and Critical Care Medicine | 6.24 | Northern America | https://www.atsjournals.org/page/ajrccm/instructions |
| 28 | Journal of Experimental Medicine | 6.24 | Northern America | https://rupress.org/jem/pages/editorial-policies#ai |

| | | | | |
|---|---|---|---|---|
| 29 | Intensive Care Medicine | 6.23 | Western Europe | https://link.springer.com/journal/134/submission-guidelines |
| 30 | Molecular Systems Biology | 6.22 | Northern America | https://www.embopress.org/page/journal/17444292/authorguide; https://www.embopress.org/page/journal/17444292/refereeguide |
| 31 | Diabetes Care | 6.01 | Northern America | https://diabetesjournals.org/care/pages/instructions-for-authors |
| 32 | Journal of Thoracic Oncology | 5.87 | Northern America | https://www.sciencedirect.com/journal/journal-of-thoracic-oncology/publish/guide-for-authors |
| 33 | European Journal of Heart Failure | 5.60 | Northern America | https://onlinelibrary.wiley.com/page/journal/18790844/homepage/forauthors.html |
| 34 | EMBO Journal | 5.48 | Western Europe | https://www.embopress.org/page/journal/14602075/authorguide; https://www.embopress.org/page/journal/14602075/authorguide#aitools; https://www.embopress.org/page/journal/14602075/refereeguide |
| 35 | Journal of the National Cancer Institute | 5.41 | Western Europe | https://academic.oup.com/jnci/pages/General_Instructions; https://publicationethics.org/cope-position-statements/ai-author |
| 36 | Journal of Clinical Investigation | 5.12 | Northern America | https://www.jci.org/kiosks/authors#COI-authors |
| 37 | Progress in Retinal and Eye Research | 4.94 | Western Europe | https://www.sciencedirect.com/journal/progress-in-retinal-and-eye-research/publish/guide-for-authors |
| 38 | Blood | 4.93 | Northern America | https://ashpublications.org/blood/pages/manuscript-prep |
| 39 | Genome Research | 4.91 | Northern America | https://genome.cshlp.org/site/misc/ifora.xhtml; https://genome.cshlp.org/site/misc/ifora_Artificial.xhtml |
| 40 | Acta Neuropathologica | 4.90 | Western Europe | https://link.springer.com/journal/401/submission-guidelines |
| 41 | Neuro-Oncology | 4.83 | Western Europe | https://academic.oup.com/neuro-oncology/pages/General_Instructions |
| 42 | Psychotherapy and Psychosomatics | 4.74 | Western Europe | https://karger.com/pps/pages/guidelines;https://karger.com/pages/peer-review#process |
| 43 | European Respiratory Journal | 4.72 | Western Europe | https://erj.ersjournals.com/authors/instructions;https://www.ersjournals.com/authors/research-ms-preparation |
| 44 | Clinical Psychology Review | 4.54 | Northern America | https://www.sciencedirect.com/journal/clinical-psychology-review/publish/guide-for-authors |
| 45 | Reports on Progress in Physics | 4.47 | Western Europe | https://publishingsupport.iopscience.iop.org/journals/reports-on-progress-in-physics/; https://publishingsupport.iopscience.iop.org/ethical-policy-journals/ |
| 46 | Journal of the American Society of Nephrology: JASN | 4.45 | Northern America | https://journals.lww.com/asnjournals/Pages/Information-for-Authors.aspx; https://journals.lww.com/asnjournals/pages/asnreviewerguidelines.aspx |
| 47 | Brain | 4.44 | Western Europe | https://academic.oup.com/brain/pages/General_Instructions |
| 48 | Drug Resistance Updates | 4.39 | Northern America | https://www.sciencedirect.com/journal/drug-resistance-updates/publish/guide-for-authors |
| 49 | eLife | 4.25 | Western Europe | https://elife-rp.msubmit.net/html/elife-rp_author_instructions.html#process |
| 50 | American Journal of Psychiatry | 4.23 | Northern America | https://ajp.psychiatryonline.org/ajp_ifora |
| 51 | PLOS Medicine | 4.22 | Northern America | https://journals.plos.org/plosmedicine/s/submission-guidelines |
| 52 | Hepatology | 4.15 | Northern America | https://onlinelibrary.wiley.com/page/journal/15273350/homepage/forauthors.html |
| 53 | Eurosurveillance | 4.15 | Western Europe | https://www.eurosurveillance.org/editorial-policy; https://www.eurosurveillance.org/for-reviewers |
| 54 | Journal of the National Comprehensive Cancer Network: JNCCN | 4.10 | Northern America | https://jnccn.org/page/forauthors/information-for-authors; https://www.icmje.org/icmje-recommendations.pdf |
| 55 | Advanced Science | 4.09 | Western Europe | https://onlinelibrary.wiley.com/page/journal/21983844/homepage/publication-ethics-guidelines |
| 56 | Radiology | 4.07 | Northern America | https://pubs.rsna.org/journal/radiology;https://pubs.rsna.org/page/policies#llm |
| 57 | Human Reproduction Update | 4.02 | Western Europe | https://academic.oup.com/humupd/pages/General |

| | | | | |
|---|---|---|---|---|
| 58 | European Journal of Epidemiology | 4.01 | Western Europe | https://www.springer.com/journal/10654/submission-guidelines |
| 59 | Protein Science | 4.01 | Northern America | https://onlinelibrary.wiley.com/page/journal/1469896x/homepage/forauthors.html |
| 60 | Clinical Infectious Diseases | 4.00 | Western Europe | https://academic.oup.com/cid/pages/Policies |
| 61 | Annals of Neurology | 3.98 | Northern America | https://onlinelibrary.wiley.com/page/journal/15318249/homepage/forauthors.html |
| 62 | Ophthalmology | 3.91 | Northern America | https://www.sciencedirect.com/journal/ophthalmology/publish/guide-for-authors |
| 63 | Journal of Infection | 3.90 | Western Europe | https://www.sciencedirect.com/journal/journal-of-infection/publish/guide-for-authors |
| 64 | Kidney International | 3.87 | Northern America | https://www.kidney-international.org/content/authorinfo |
| 65 | Nano Today | 3.87 | Western Europe | https://www.sciencedirect.com/journal/nano-today/publish/guide-for-authors |
| 66 | Annals of Internal Medicine | 3.85 | Northern America | https://www.acpjournals.org/journal/aim/authors |
| 67 | Arthritis and Rheumatology | 3.79 | Western Europe | https://acrjournals.onlinelibrary.wiley.com/hub/journal/23265205/forauthors.html; https://authorservices.wiley.com/ethics-guidelines/index.html |
| 68 | Journal of Allergy and Clinical Immunology | 3.74 | Northern America | https://www.jacionline.org/content/authorinfo |
| 69 | Immunological Reviews | 3.73 | Western Europe | https://onlinelibrary.wiley.com/page/journal/1600065x/homepage/forauthors.html |
| 70 | GigaScience | 3.70 | Western Europe | https://academic.oup.com/gigascience/pages/instructions_to_authors |
| 71 | Journal of Cell Biology | 3.66 | Northern America | https://rupress.org/jcb/pages/ifora;https://rupress.org/jcb/pages/editorial-policies#ai |
| 72 | Journal of the American Academy of Child and Adolescent Psychiatry | 3.66 | Western Europe | https://www.jaacap.org/content/authorinfo |
| 73 | Cancer Treatment Reviews | 3.59 | Western Europe | https://www.cancertreatmentreviews.com/content/authorinfo |
| 74 | Clinical Microbiology and Infection | 3.59 | Western Europe | https://www.clinicalmicrobiologyandinfection.com/content/authorinfo |
| 75 | Journal of Extracellular Vesicles | 3.53 | Northern America | https://onlinelibrary.wiley.com/page/journal/20013078/homepage/author-guidelines |
| 76 | Health Affairs | 3.50 | Northern America | https://www.healthaffairs.org/help-for-authors/policies |
| 77 | Small | 3.40 | Western Europe | https://onlinelibrary.wiley.com/page/journal/16136829/homepage/author-guidelines; https://onlinelibrary.wiley.com/page/journal/16136829/homepage/reviewer-guidelines |
| 78 | Experimental and Molecular Medicine | 3.38 | Asiatic Region | https://www.nature.com/documents/EMM_GTA.pdf |
| 79 | Protein and Cell | 3.37 | Asiatic Region | https://www.springer.com/journal/13238/submission-guidelines |
| 80 | American Psychologist | 3.36 | Northern America | https://www.apa.org/pubs/journals/amp/ |
| 81 | Journal of Clinical Epidemiology | 3.36 | Northern America | https://www.jclinepi.com/content/authorinfo |
| 82 | Diabetologia | 3.35 | Western Europe | https://diabetologia-journal.org/for-authors-and-reviewers/instructions-to-authors/ |
| 83 | Sports Medicine | 3.29 | Western Europe | https://link.springer.com/journal/40279/submission-guidelines |
| 84 | Alzheimer’s and Dementia | 3.29 | Northern America | https://www.keaipublishing.com/en/journals/engineered-regeneration/guide-for-authors/ |
| 85 | Engineered Regeneration | 3.25 | Asiatic Region | https://www.keaipublishing.com/en/authors-and-editors/publishing-ethics/ |
| 86 | FEMS Microbiology Reviews | 3.25 | Western Europe | https://academic.oup.com/femsre/pages/Manuscript_Preparation |
| 87 | Molecular Aspects of Medicine | 3.24 | Western Europe | https://www.sciencedirect.com/journal/molecular-aspects-of-medicine/publish/guide-for-authors |
| 88 | Medical Image Analysis | 3.20 | Western Europe | https://www.sciencedirect.com/journal/medical-image-analysis/publish/guide-for-authors |
| 89 | Health Psychology Review | 3.10 | Western Europe | https://www.tandfonline.com/action/authorSubmission?show=instructions&journalCode=rhpr20 |

| | | | | |
|---|---|---|---|---|
| 90 | Journal of Thrombosis and Haemostasis | 3.09 | Western Europe | https://www.sciencedirect.com/journal/journal-of-thrombosis-and-haemostasis/publish/guide-for-authors |
| 91 | American Journal of Obstetrics and Gynecology | 3.07 | Northern America | https://www.sciencedirect.com/journal/american-journal-of-obstetrics-and-gynecology/publish/guide-for-authors |
| 92 | Pharmacology and Therapeutics | 3.06 | Northern America | https://www.sciencedirect.com/journal/pharmacology-and-therapeutics/publish/guide-for-authors |
| 93 | Autism in Adulthood | 3.06 | Northern America | https://home.liebertpub.com/publications/646/pdf |
| 94 | Sleep Medicine Reviews | 3.05 | Western Europe | https://www.sciencedirect.com/journal/sleep-medicine-reviews/publish/guide-for-authors |
| 95 | Journal of Child Psychology and Psychiatry and Allied Disciplines | 3.03 | Western Europe | https://acamh.onlinelibrary.wiley.com/hub/journal/14697610/forauthors.html; https://authorservices.wiley.com/Reviewers/index.html |
| 96 | Journal of Internal Medicine | 2.99 | Western Europe | https://onlinelibrary.wiley.com/page/journal/13652796/homepage/forauthors.html |
| 97 | American Journal of Kidney Diseases | 2.97 | Western Europe | https://sites.google.com/site/ajkdinfoforauthors/#h.p_fuqD1SgJB-N0 |
| 98 | Liver Cancer | 2.96 | Western Europe | https://karger.com/lic/pages/guidelines |

**Supplementary Table 2. Journal characteristics and information sources of GAI usage guidelines of whole-spectrum sample journals**

| ID | Journal name | SJR Score | Region | Website |
|---|---|---|---|---|
| 1 | Genome Medicine | 4.85 | Western Europe | https://www.biomedcentral.com/getpublished/editorial-policies#artificial+intelligence+%28ai%29 |
| 2 | JAMA Neurology | 6.70 | Northern America | https://jamanetwork.com/journals/jamaneurology/pages/instructions-for-authors |
| 3 | Cancer Treatment Reviews | 3.59 | Western Europe | https://www.sciencedirect.com/journal/cancer-treatment-reviews/publish/guide-for-authors |
| 4 | Journal of Extracellular Vesicles | 3.53 | Northern America | https://onlinelibrary.wiley.com/page/journal/20013078/homepage/author-guidelines |
| 5 | Alimentary Pharmacology and Therapeutics | 2.79 | Western Europe | https://onlinelibrary.wiley.com/page/journal/13652036/homepage/forauthors.html |
| 6 | JACC: Clinical Electrophysiology | 2.56 | Northern America | https://www.sciencedirect.com/journal/jacc-clinical-electrophysiology/publish/guide-for-authors |
| 7 | JNCI Cancer Spectrum | 2.38 | Northern America | https://academic.oup.com/jncics/pages/General_Instructions |
| 8 | Cancer Letters | 2.09 | Western Europe | https://www.sciencedirect.com/journal/cancer-letters/publish/guide-for-authors |
| 9 | Obesity | 1.83 | Northern America | https://onlinelibrary.wiley.com/page/journal/1930739x/homepage/forauthors.html |
| 10 | Reviews in Endocrine and Metabolic Disorders | 1.84 | Western Europe | https://link.springer.com/journal/11154/submission-guidelines |
| 11 | Medicine and Science in Sports and Exercise | 1.73 | Northern America | https://edmgr.ovid.com/msse/accounts/ifauth.htm |
| 12 | Vaccines | 1.66 | Western Europe | https://www.sciencedirect.com/journal/vaccine/publish/guide-for-authors |
| 13 | Journal of Headache and Pain | 1.59 | Western Europe | https://www.biomedcentral.com/getpublished/editorial-policies#artificial+intelligence+%28ai%29 |
| 14 | General Psychiatry | 1.50 | Western Europe | https://gpsych.bmj.com/pages/authors#submission_guidelines |
| 15 | Best Practice and Research in Clinical Endocrinology and Metabolism | 1.43 | Western Europe | https://www.sciencedirect.com/journal/best-practice-and-research-clinical-endocrinology-and-metabolism/publish/guide-for-authors |
| 16 | ACS Synthetic Biology | 1.41 | Northern America | https://publish.acs.org/publish/author_guidelines?coden=asbcd6 |
| 17 | International Journal of Sports Physiology and Performance | 1.33 | Northern America | https://journals.humankinetics.com/view/journals/ijspp/ijspp-overview.xml?tab_body=author-guidelines |
| 18 | eNeuro | 1.31 | Northern America | https://www.eneuro.org/content/general-information#policies |
| 19 | Science and Medicine in Football | 1.23 | Western Europe | https://www.tandfonline.com/action/authorSubmission?show=instructions&journalCode=rsmf20 |
| 20 | Journal of Endocrinology | 1.28 | Western Europe | https://joe.bioscientifica.com/page/107 |
| 21 | International Journal of Health Policy and Management | 1.16 | Middle East | https://www.ijhpm.com/journal/authors.note |
| 22 | Archives of Toxicology | 1.16 | Western Europe | https://link.springer.com/journal/204/submission-guidelines |
| 23 | International Journal of Nanomedicine | 1.11 | Pacific Region | https://www.tandfonline.com/action/authorSubmission?show=instructions&journalCode=dijn20 |
| 24 | Journal of Studies on Alcohol and Drugs | 1.12 | Northern America | https://www.jsad.com/page/instructions |
| 25 | American Journal of Drug and Alcohol Abuse | 1.07 | Northern America | https://www.tandfonline.com/action/authorSubmission?show=instructions&journalCode=iada20 |
| 26 | Biomedical Signal Processing and Control | 1.07 | Western Europe | https://www.sciencedirect.com/journal/biomedical-signal-processing-and-control/publish/guide-for-authors |
| 27 | Journal of Lipid and Atherosclerosis | 1.03 | Asiatic Region | https://e-jla.org/index.php?body=instructions |

| | | | | |
|---|---|---|---|---|
| 28 | Journal of Pediatrics | 1.04 | Northern America | https://www.jpeds.com/content/authorinfo |
| 29 | Cancer Biology and Therapy | 1.00 | Northern America | https://www.tandfonline.com/action/authorSubmission?show=instructions&journalCode=kcbt20 |
| 30 | Journal of Huntington's disease | 1.02 | Western Europe | https://www.iospress.com/catalog/journals/journal-of-huntingtons-disease |
| 31 | Clinical and Experimental Rheumatology | 0.98 | Western Europe | https://www.clinexprheumatol.org/guidelines-authors.asp |
| 32 | Nanotheranostics | 0.95 | Pacific Region | https://www.ntno.org/ms/author |
| 33 | International Journal of Oral Implantology | 0.94 | Northern America | https://www.quintpub.com/journals/ejoi/gp.php?journal_name=EJOI&name_abbr=EJOI; file:///D:/0.2023_Fall/conference%20&%20research/guideline%20data%202%20PDF/033-authorguidelines_ejoi.pdf |
| 34 | Journal of Diabetes Research | 0.93 | Africa/Middle East | https://www.hindawi.com/journals/jdr/guidelines/ |
| 35 | Microorganisms | 0.91 | Western Europe | https://www.mdpi.com/journal/microorganisms/instructions |
| 36 | Archives of Environmental Contamination and Toxicology | 0.89 | Northern America | https://link.springer.com/journal/244/submission-guidelines |
| 37 | Journal of Reproductive and Infant Psychology | 0.87 | Western Europe | https://www.tandfonline.com/action/authorSubmission?show=instructions&journalCode=cjri20 |
| 38 | Osong Public Health and Research Perspectives | 0.86 | Asiatic Region | https://www.ophrp.org/authors/authors.php |
| 39 | European Journal of Anaesthesiology | 0.85 | Western Europe | https://edmgr.ovid.com/eja/accounts/ifauth.htm |
| 40 | Asian Pacific Journal of Allergy and Immunology | 0.85 | Asiatic Region | https://apjai-journal.org/wp-content/uploads/2023/11/author-guidelines-for-APJAI_Nov2023.pdf |
| 41 | Bioinspiration and Biomimetics | 0.81 | Western Europe | https://publishingsupport.iopscience.iop.org/journals/bioinspiration-biomimetics/ |
| 42 | Journal of Immunological Methods | 0.83 | Western Europe | https://www.sciencedirect.com/journal/journal-of-immunological-methods/publish/guide-for-authors |
| 43 | Topics in antiviral medicine | 0.78 | Northern America | https://www.iasusa.org/wp-content/uploads/guidelines/arv/2020-Guidelines-for-Author-and-Contributors.pdf |
| 44 | Advances in Radiation Oncology | 0.79 | Northern America | https://www.astro.org/News-and-Publications/Journals/Author-Instructions |
| 45 | International Journal of Occupational and Environmental Medicine | 0.76 | Middle East | http://ijomeh.eu/Instructions-for-Authors,120.html |
| 46 | Clinical Psychology in Europe | 0.76 | Western Europe | https://cpe.psychopen.eu/index.php/cpe/policies |
| 47 | American Journal of Perinatology | 0.73 | Northern America | https://lp.thieme.de/open-access-files/113/author_instructions.pdf |
| 48 | World Journal of Microbiology and Biotechnology | 0.73 | Western Europe | https://link.springer.com/journal/11274/submission-guidelines |
| 49 | Archives of Osteoporosis | 0.72 | Western Europe | file:///D:/0.2023_Fall/conference%20&%20research/guideline%20data%202%20PDF/048-OI%20Instructions%20for%20Authors_August_2023.pdf |
| 50 | Infectious Diseases Now | 0.71 | Western Europe | https://www.sciencedirect.com/journal/infectious-diseases-now/publish/guide-for-authors |
| 51 | Drug Target Insights | 0.69 | Western Europe | https://us.sagepub.com/en-us/nam/manuscript-submission-guidelines |
| 52 | Journal of Biomechanics | 0.70 | Western Europe | https://www.sciencedirect.com/journal/journal-of-biomechanics/publish/guide-for-authors |

| | | | | |
|---|---|---|---|---|
| 53 | Substance Abuse: Research and Treatment | 0.68 | Northern America | https://us.sagepub.com/en-us/nam/substance-abuse-research-and-treatment/journal202697#submission-guidelines |
| 54 | Journal of Medical Microbiology | 0.67 | Western Europe | https://www.microbiologyresearch.org/ethics-policies |
| 55 | Journal of Cancer Education | 0.65 | Northern America | https://link.springer.com/journal/13187/submission-guidelines |
| 56 | Open Access Rheumatology: Research and Reviews | 0.66 | Pacific Region | https://www.tandfonline.com/action/authorSubmission?show=instructions&journalCode=doar20 |
| 57 | American Journal of Psychoanalysis | 0.64 | Western Europe | https://amjpa.org/about-the-ajp/instructions-for-authors/#1523389239352-806955fc-7d21 |
| 58 | Global Health Promotion | 0.64 | Western Europe | https://journals.sagepub.com/pb-assets/cmscontent/ped/GHP%20Peer%20Review%20English%20Guidelines-temporary-website-update8NOv2022a-1668748975.pdf |
| 59 | Future Science OA | 0.62 | Western Europe | https://www.future-science.com/authorguide/editorialpolicies |
| 60 | Gynecologic and Obstetric Investigation | 0.61 | Western Europe | https://karger.com/goi/pages/guidelines |
| 61 | Iranian Journal of Psychiatry | 0.58 | Middle East | https://ijps.tums.ac.ir/;https://ijps.tums.ac.ir/index.php/ijps/peer_review_policy |
| 62 | Archives of Virology | 0.60 | Western Europe | https://www.springer.com/journal/705/submission-guidelines |
| 63 | Emergencias | 0.57 | Western Europe | https://emergencias.portalsemes.org/informacion-para-autores/english/;https://emergencias.portalsemes.org/images/info-reviewers-en.pdf |
| 64 | American Family Physician | 0.57 | Northern America | https://www.aafp.org/pubs/afp/authors.html |
| 65 | AIMS Public Health | 0.56 | Northern America | https://www.aimspress.com/aimsph/news/solo-detail/instructionsforauthors; https://www.aimspress.com/aimsph/news/solo-detail/peerreviewguidelines |
| 66 | Photodermatology Photoimmunology and Photomedicine | 0.56 | Western Europe | https://onlinelibrary.wiley.com/page/journal/16000781/homepage/forauthors.html |
| 67 | World Journal of Orthopedics | 0.53 | Asiatic Region | https://www.wjgnet.com/bpg/gerinfo/204 |
| 68 | Archives of Insect Biochemistry and Physiology | 0.53 | Western Europe | https://onlinelibrary.wiley.com/page/journal/15206327/homepage/forauthors.html |
| 69 | JMIR Research Protocols | 0.53 | Northern America | https://www.researchprotocols.org/author-information/instructions-for-authors |
| 70 | Orthopedics | 0.52 | Northern America | https://journals.healio.com/journal/ortho/submit-an-article#Authors |
| 71 | Asian Journal of Surgery | 0.51 | Asiatic Region | https://www.sciencedirect.com/journal/asian-journal-of-surgery/publish/guide-for-authors |
| 72 | Spine Surgery and Related Research | 0.50 | Asiatic Region | http://ssrr-journal.jp/authors/ |
| 73 | Journal of Neurosurgical Sciences | 0.49 | Western Europe | https://www.minervamedica.it/en/journals/neurosurgical-sciences/notice-to-authors.php |
| 74 | Current Molecular Medicine | 0.49 | Western Europe | https://benthamscience.com/pages/author-guidelines |
| 75 | Advances in Urology | 0.47 | Northern America | https://www.hindawi.com/journals/au/guidelines/; https://www.hindawi.com/publish-research/reviewers/ |
| 76 | Communication in Biomathematical Sciences | 0.47 | Asiatic Region | https://journals.itb.ac.id/index.php/cbms/guideline |
| 77 | Acta Pharmaceutica | 0.46 | Eastern Europe | https://acta.pharmaceutica.farmaceut.org/instructions-to-authors/ |
| 78 | Pleura and Peritoneum | 0.45 | Western Europe | https://www.degruyter.com/journal/key/pp/html?lang=en#submit |
| 79 | Innovative Surgical Sciences | 0.43 | Western Europe | https://www.degruyter.com/journal/key/iss/html?lang=en#submit |

| | | | | |
|---|---|---|---|---|
| 80 | Journal of Pediatric Pharmacology and Therapeutics | 0.44 | Northern America | https://meridian.allenpress.com/DocumentLibrary/PPAG/JPPT_Instruction_for_Authors-12-3-2020.pdf |
| 81 | Comparative Medicine | 0.41 | Northern America | https://www.aalas.org/publications/information-for-authors/cm-and-jaalas/manuscript-preparation |
| 82 | Journal of Education and Health Promotion | 0.41 | Asiatic Region | https://journals.lww.com/JEHP/Pages/informationforauthors.aspx |
| 83 | Journal of Military and Veterans' Health | 0.40 | Pacific Region | https://jmvh.org/authors/;https://jmvh.org/reviewers/ |
| 84 | Journal of Bodywork and Movement Therapies | 0.40 | Northern America | https://www.bodyworkmovementtherapies.com/content/authorinfo |
| 85 | Jornal Brasileiro de Reproducao Assistida | 0.38 | Latin America | https://www.jbra.com.br/mensagem/pub/mensagem.php?id_mensagem=custom_sites&lingua_atual=_ing |
| 86 | Cambridge Quarterly of Healthcare Ethics | 0.39 | Western Europe | https://www.cambridge.org/core/journals/cambridge-quarterly-of-healthcare-ethics/information/author-instructions/preparing-your-materials |
| 87 | Folia Histochemica et Cytobiologica | 0.37 | Eastern Europe | https://journals.viamedica.pl/folia_histochemica_cytobiologica/about/submissions#authorGuidelines |
| 88 | Frontiers in Bioscience - Scholar | 0.37 | Asiatic Region | https://www.imrpress.com/journal/FBS/instructions |
| 89 | Acta Cardiologica | 0.36 | Western Europe | https://www.tandfonline.com/toc/tacd20/current; https://www.tandfonline.com/action/authorSubmission?show=instructions&journalCode=tacd20 |
| 90 | Annals of Transplantation | 0.35 | Northern America | https://annalsoftransplantation.com/;https://annalsoftransplantation.com/instructions |
| 91 | Iranian Journal of Pharmaceutical Research | 0.34 | Middle East | https://brieflands.com/journals/iranian-journal-of-pharmaceutical-research/; https://brieflands.com/journals/iranian-journal-of-pharmaceutical-research/knowledgebase/category/tree |
| 92 | Acta Microbiologica et Immunologica Hungarica | 0.33 | Eastern Europe | https://akjournals.com/view/journals/030/030-overview.xml?rskey=k4UGYp&result=17 |
| 93 | Journal of dental hygiene : JDH / American Dental Hygienists' Association | 0.32 | Northern America | https://jdh.adha.org/ |
| 94 | Indian Journal of Traditional Knowledge | 0.31 | Asiatic Region | http://op.niscair.res.in/index.php/IJTK |
| 95 | Revista Brasileira de Ginecologia e Obstetricia | 0.30 | Latin America | https://www.scielo.br/j/rbgo/ |
| 96 | Journal of Opioid Management | 0.30 | Northern America | https://wmpllc.org/ojs/index.php/jom |
| 97 | Tropical Life Sciences Research | 0.29 | Asiatic Region | https://ejournal.usm.my/tlsr |
| 98 | American Society of Clinical Oncology educational book / ASCO. American Society of Clinical Oncology. Meeting | 0.29 | Northern America | https://ascopubs.org/journal/edbk |
| 99 | Acta Dermatovenerologica Alpina. Panonica et Adriatica | 0.27 | Eastern Europe | https://acta-apa.org/ |
| 100 | Anales de Pediatria | 0.27 | Western Europe | https://analesdepediatria.org/en |

| | | | | |
|---|---|---|---|---|
| 101 | Cardiovascular Journal of Africa | 0.26 | Africa | https://www.cvja.co.za/ |
| 102 | Experimental and Clinical Transplantation | 0.26 | Middle East | https://www.ectrx.org/ |
| 103 | Pediatric Medicine | 0.24 | Asiatic Region | https://pm.amegroups.org/ |
| 104 | Journal of clinical orthodontics : JCO | 0.24 | Northern America | https://www.jco-online.com/ |
| 105 | Acta Orthopaedica Belgica | 0.22 | Western Europe | http://www.actaorthopaedica.be/ |
| 106 | Romanian Journal of Morphology and Embryology | 0.23 | Eastern Europe | https://rjme.ro/ |
| 107 | Journal of Allied Health | 0.22 | Northern America | https://www.asahp.org/journal-of-allied-health |
| 108 | Tanaffos | 0.21 | Middle East | https://www.tanaffosjournal.ir/ |
| 109 | Vestnik Otorinolaringologii | 0.21 | Eastern Europe | https://www.mediasphera.ru/journal/vestnik-otorinolaringologii?clear_cache=Y |
| 110 | Doklady Biochemistry and Biophysics | 0.20 | Northern America | https://link.springer.com/journal/10628 |
| 111 | Asia Pacific Journal of Health Management | 0.19 | Pacific Region | https://journal.achsm.org.au/index.php/achsm |
| 112 | Journal of Medical Regulation | 0.19 | Northern America | https://meridian.allenpress.com/jmr/pages/For-Authors |
| 113 | Journal of Oral and Maxillofacial Surgery. Medicine. and Pathology | Elsevier Ltd. | United Kingdom | https://www.sciencedirect.com/journal/journal-of-oral-and-maxillofacial-surgery-medicine-and-pathology/publish/guide-for-authors |
| 114 | Palliative Medicine in Practice | 0.18 | Eastern Europe | https://journals.viamedica.pl/palliative_medicine_in_practice |
| 115 | Trauma | 0.17 | Western Europe | https://journals.sagepub.com/home/tra |
| 116 | Wounds UK | 0.17 | Western Europe | https://wounds-uk.com/journal-articles/wounds-uk-2018-award-winners/ |
| 117 | Medical Journal of Indonesia | 0.17 | Asiatic Region | https://mji.ui.ac.id/journal/index.php/mji/ifa |
| 118 | Academic Forensic Pathology | 0.16 | Western Europe | https://journals.sagepub.com/author-instructions/AFP |
| 119 | B-ENT | 0.15 | Western Europe | http://www.b-ent.be/en/instructions-to-authors-106 |
| 120 | South Eastern European Journal of Public Health | 0.15 | Asiatic Region | https://www.seejph.com/index.php/seejph/about/submissions |
| 121 | Pakistan Journal of Life and Social Sciences | 0.15 | Asiatic Region | https://www.pjlss.edu.pk/instructions.htm |
| 122 | Journal of Sichuan University (Medical Science Edition) | 0.14 | Asiatic Region | http://jsu-mse.com/index.php/journal/pages/view/authorguidelines |
| 123 | Formosan Journal of Surgery | 0.14 | Asiatic Region | https://edmgr.ovid.com/fjs/accounts/ifauth.htm |
| 124 | Malaysian Journal of Microbiology | 0.13 | Northern America | http://mjm.usm.my/index.php?r=cms/entry/view&id=64&slug=Preparation-of-manuscript |
| 125 | South Dakota journal of medicine | 0.13 | Africa | https://www.sdsma.org/AuthorSubmit |
| 126 | African Journal Biomedical Research | 0.13 | Pacific Region | https://www.ajol.info/index.php/ajbr/about/submissions |
| 127 | Immunology and Immunogenetics Insights | 0.13 | Northern America | https://journals.sagepub.com/home/iii; https://us.sagepub.com/en-us/nam/manuscript-submission-guidelines |

| | | | | |
|---|---|---|---|---|
| 128 | Analytical and Quantitative Cytopathology and Histopathology | 0.13 | Asiatic Region | https://www.aqch.com/; https://www.aqch.com/AQCH_AuthorChecklist.pdf |
| 129 | African Journal of Paediatric Surgery | 0.13 | Asiatic Region | https://journals.lww.com/AJPS/Pages/informationforauthors.aspx |
| 130 | Journal of Clinical Hepatology | 0.12 | Eastern Europe | http://www.lcgdbzz.org/news/EditorialPolicies.htm |
| 131 | Siberian Journal of Oncology | 0.12 | Eastern Europe | https://www.siboncoj.ru/jour/about/submissions#authorGuidelines |
| 132 | Antibiotiki i Khimioterapiya | 0.12 | Middle East | https://www.antibiotics-chemotherapy.ru/jour/about/submissions#authorGuidelines |
| 133 | Studia Universitatis Vasile Goldis Arad. Seria Stiintele Vietii | 0.12 | Eastern Europe | http://www.studiauniversitatis.ro/publication-ethics/ |
| 134 | International Journal of Medical Toxicology and Forensic Medicine | 0.11 | Middle East | https://journals.sbmu.ac.ir/ijmtfm/author_guidelines |
| 135 | Anatolian Journal of Family Medicine | 0.11 | Middle East | https://ajfamed.org/policies |
| 136 | Mediterranean Journal of Infection. Microbes and Antimicrobials | 0.11 | Asiatic Region | https://mjima.org/static.php?id=4 |
| 137 | International Eye Science | 0.11 | Northern America | http://www.ijo.cn/gjyken/site/menus/20220929162530001 |
| 138 | Vascular Disease Management | 0.11 | Eastern Europe | https://www.hmpgloballearningnetwork.com/site/vdm/instructions-for-authors |
| 139 | Turkish Journal of Plastic Surgery | 0.11 | Asiatic Region | https://journals.lww.com/tjps/Pages/informationforauthors.aspx |
| 140 | Paediatria Croatica | 0.10 | Eastern Europe | https://www.paedcro.com/en/authors |
| 141 | Current Gynecologic Oncology | 0.10 | Western Europe | http://www.ginekologia.pl/index.php/instructions-for-authors |
| 142 | Forum of Clinical Oncology | 0.10 | Eastern Europe | https://sciendo-parsed-data-feed.s3.eu-central-1.amazonaws.com/FCO/Author_Guidelines.pdf |
| 143 | New Medicine | 0.10 | Western Europe | http://www.newmedicine.pl/guide/ |
| 144 | Technische Sicherheit | 0.10 | Western Europe | https://technikwissen.eu/for-authors/ |

**Supplementary Table 3. Provision of specific requirements of different GAI usage guidelines**

| | Top journals (n = 98) | Whole-spectrum sample journals (n = 144) | P value* |
|---|---|---|---|
| **Author guidelines, n (%)** | | | |
| Usage permission | 63 (64.3) | 40 (27.8) | < 0.01 |
| Language editing | 47 (48.0) | 22 (15.3) | < 0.01 |
| Manuscript writing | 48 (49.0) | 23 (16.0) | < 0.01 |
| Data analysis and interpretation | 39 (39.8) | 18 (12.5) | < 0.01 |
| Image generating | 39 (39.8) | 22 (15.3) | < 0.01 |
| Fact-checking requirement | 44 (44.9) | 22 (15.3) | < 0.01 |
| Usage documentation | 63 (64.3) | 38 (26.4) | < 0.01 |
| Authorship eligibility | 62 (63.3) | 38 (26.4) | < 0.01 |
| **Reviewer guidelines, n (%)** | | | |
| Usage permission | 11 (11.2) | 0 (0.0) | < 0.01 |
| Language editing | 9 (9.2) | 0 (0.0) | < 0.01 |
| Usage documentation | 5 (5.1) | 0 (0.0) | < 0.01 |
| **Specificity score, mean (SD)** | | | |
| Author guidelines (range: 0-8) | 4.1 (3.4) | 1.6 (2.8) | < 0.01 |
| Reviewer guidelines (range: 0-3) | 0.3 (0.7) | 0.0 (0.0) | < 0.01 |
| Author and reviewer guidelines (range: 0-11) | 4.4 (3.7) | 1.6 (2.8) | < 0.01 |
| **Reference to external guidelines, n (%)** | | | |
| COPE | 49 (50.0) | 61 (42.4) | 0.30 |
| ICMJE | 54 (55.1) | 82 (57.0) | 0.88 |
| WAME | 6 (6.1) | 12 (8.3) | 0.70 |
| Publishers | 49 (50.0) | 18 (12.5) | < 0.01 |

Footnote: * Differences between the two groups of journals were analyzed using Wilcoxon Rank Sum tests or Chi-Square tests.

**Supplementary Table 4. External GAI usage guidelines and their requirements**

| Guidelines on GAI usage for authors and/or reviewers | |
|---|---|
| **Committee on Publication Ethics (COPE)** | The use of artificial intelligence (AI) tools such as ChatGPT or Large Language Models in research publications is expanding rapidly. COPE joins organizations, such as WAME and the JAMA Network among others, to state that AI tools cannot be listed as an author of a paper. AI tools cannot meet the requirements for authorship as they cannot take responsibility for the submitted work. As non-legal entities, they cannot assert the presence or absence of conflicts of interest nor manage copyright and license agreements. Authors who use AI tools in the writing of a manuscript, production of images or graphical elements of the paper, or in the collection and analysis of data, must be transparent in disclosing in the Materials and Methods (or similar section) of the paper how the AI tool was used and which tool was used. Authors are fully responsible for the content of their manuscript, even those parts produced by an AI tool, and are thus liable for any breach of publication ethics.<br><br>AI powered automation to increase processing speed, validation, quality assessment, and progression of the peer review process can be used, is acceptable, and even expected in many cases, provided the outcome does not result in a decision by the AI itself on acceptance or rejection of a manuscript. For example, if an AI tool detects a figure in a manuscript including a recognisable human face without the required consent form, the issue should be raised to the attention of the editor to make a decision on rejection, or the automation could proceed to automatically send a message requesting clarification, or the relevant documentation, from the authors. Publishers should take steps to be transparent about which of their publishing processes or workflows are automated, and where AI decisions are involved. Any AI powered automation should be clearly presented to the relevant participants of the peer review process—authors, reviewers, or editors—with clarification on how the algorithm provided the result or conclusion. |
| **International Committee of Medical Journal Editors (ICMJE)** | At submission, the journal should require authors to disclose whether they used artificial intelligence (AI)-assisted technologies (such as Large Language Models [LLMs], chatbots, or image creators) in the production of submitted work. Authors who use such technology should describe, in both the cover letter and the submitted work, how they used it. Chatbots (such as ChatGPT) should not be listed as authors because they cannot be responsible for the accuracy, integrity, and originality of the work, and these responsibilities are required for authorship (see Section II.A.1). Therefore, humans are responsible for any submitted material that included the use of AI-assisted technologies. Authors should carefully review and edit the result because AI can generate authoritative-sounding output that can be incorrect, incomplete, or biased. Authors should not list AI and AI-assisted technologies as an author or co-author, nor cite AI as an author. Authors should be able to assert that there is no plagiarism in their paper, including in text and images produced by the AI. Humans must ensure there is appropriate attribution of all quoted material, including full citations.<br><br>Peer Reviewers Manuscripts submitted to journals are privileged communications that are authors' private, confidential property, and authors may be harmed by premature disclosure of any or all of a manuscript's details. Reviewers therefore should keep manuscripts and the information they contain strictly confidential. Reviewers must not publicly discuss authors' work and must not appropriate authors' ideas before the manuscript is published. Reviewers must not retain the manuscript for their personal use and should destroy copies of manuscripts after submitting their reviews. Reviewers who seek assistance from a trainee or colleague in the performance of a review should acknowledge these individuals' contributions in the written comments submitted to the editor. Reviewers must maintain the confidentiality of the manuscript as outlined above, which may prohibit the uploading of the manuscript to software or other AI technologies where confidentiality cannot be assured. Reviewers should disclose to journals if and how AI technology is being used to facilitate their review. Reviewers should be aware that AI can generate authoritative-sounding output that can be incorrect, incomplete, or biased. Reviewers are expected to respond promptly to |

| | |
|---|---|
| | requests to review and to submit reviews within the time agreed. Reviewers' comments should be constructive, honest, and polite. Reviewers should declare their relationships and activities that might bias their evaluation of a manuscript and recuse themselves from the peer-review process if a conflict exists. |
| **World Association of Medical Editors (WAME)** | Chatbots cannot be authors. Journals have begun to publish articles in which chatbots such as Bard, Bing and ChatGPT have been used, with some journals listing chatbots as co-authors. The legal status of an author differs from country to country but under most jurisdictions, an author must be a legal person. Chatbots do not meet the International Committee of Medical Journal Editors (ICMJE) authorship criteria, particularly that of being able to give "final approval of the version to be published" and "to be accountable for all aspects of the work in ensuring that questions related to the accuracy or integrity of any part of the work are appropriately investigated and resolved." (10) No AI tool can "understand" a conflict-of-interest statement, and does not have the legal standing to sign a statement. Chatbots have no affiliation independent of their developers. Since authors submitting a manuscript must ensure that all those named as authors meet the authorship criteria, chatbots cannot be included as authors.<br>Authors should be transparent when chatbots are used and provide information about how they were used. The extent and type of use of chatbots in journal publications should be indicated. This is consistent with the ICMJE recommendation of acknowledging writing assistance (11) and providing in the Methods detailed information about how the study was conducted and the results generated. (12) Authors submitting a paper in which a chatbot/AI was used to draft new text should note such use in the acknowledgment; all prompts used to generate new text, or to convert text or text prompts into tables or illustrations, should be specified. When an AI tool such as a chatbot is used to carry out or generate analytical work, help report results (e.g., generating tables or figures), or write computer codes, this should be stated in the body of the paper, in both the Abstract and the Methods section. In the interests of enabling scientific scrutiny, including replication and identifying falsification, the full prompt used to generate the research results, the time and date of query, and the AI tool used and its version, should be provided.<br>Authors are responsible for material provided by a chatbot in their paper (including the accuracy of what is presented and the absence of plagiarism) and for appropriate attribution of all sources (including original sources for material generated by the chatbot). Authors of articles written with the help of a chatbot are responsible for the material generated by the chatbot, including its accuracy. Noting that plagiarism is "the practice of taking someone else's work or ideas and passing them off as one's own" (13), not just the verbatim repetition of previously published text. It is the author's responsibility to ensure that the content reflects the author's data and ideas and is not plagiarism, fabrication or falsification. Otherwise, it is potentially scientific misconduct to offer such material for publication, irrespective of how it was written. Similarly, authors must ensure that all quoted material is appropriately attributed, including full citations, and that the cited sources support the chatbot's statements. Since a chatbot may be designed to omit sources that oppose viewpoints expressed in its output, it is the authors' responsibility to find, review and include such counterviews in their articles. (Of course, such biases are also found in human authors.) Authors should identify the chatbot used and the specific prompt (query statement) used with the chatbot. They should specify what they have done to mitigate the risk of plagiarism, provide a balanced view, and ensure the accuracy of all their references.<br><br>Reviewer guidelines: Editors and peer reviewers should specify, to authors and each other, any use of chatbots in the evaluation of the manuscript and generation of reviews and correspondence. If they use chatbots in their communications with authors and each other, they should explain how they were used. Editors and reviewers are responsible for any content and citations generated by a chatbot. They should be aware that chatbots retain the prompts fed to them, including manuscript content, and supplying an author's manuscript to a chatbot breaches confidentiality of the submitted manuscript. |

| | |
|---|---|
| **Elsevier (Publisher)** | The use of generative AI and AI-assisted technologies in scientific writing This policy has been triggered by the rise of generative AI and AI-assisted technologies which are expected to increasingly be used by content creators. The policy aims to provide greater transparency and guidance to authors, readers, reviewers, editors and contributors. Elsevier will monitor this development and will adjust or refine this policy when appropriate. Please note the policy only refers to the writing process, and not to the use of AI tools to analyse and draw insights from data as part of the research process. Where authors use generative AI and AI-assisted technologies in the writing process, these technologies should only be used to improve readability and language of the work. Applying the technology should be done with human oversight and control and authors should carefully review and edit the result, because AI can generate authoritative-sounding output that can be incorrect, incomplete or biased. The authors are ultimately responsible and accountable for the contents of the work. Authors should disclose in their manuscript the use of AI and AI-assisted technologies and a statement will appear in the published work. Declaring the use of these technologies supports transparency and trust between authors, readers, reviewers, editors and contributors and facilitates compliance with the terms of use of the relevant tool or technology. Authors should not list AI and AI-assisted technologies as an author or co-author, nor cite AI as an author. Authorship implies responsibilities and tasks that can only be attributed to and performed by humans. Each (co-) author is accountable for ensuring that questions related to the accuracy or integrity of any part of the work are appropriately investigated and resolved and authorship requires the ability to approve the final version of the work and agree to its submission. Authors are also responsible for ensuring that the work is original, that the stated authors qualify for authorship, and the work does not infringe third party rights, and should familiarize themselves with our Ethics in Publishing policy before they submit.<br><br>This policy has been triggered by the rise of generative AI and AI-assisted technologies* and aims to provide greater transparency and guidance to authors, editors and reviewers. Elsevier will monitor ongoing developments in this area closely and will adjust or refine the policy as appropriate. The following guidance is specifically for reviewers. When a researcher is invited to review another researcher's paper, the manuscript must be treated as a confidential document. Reviewers should not upload a submitted manuscript or any part of it into a generative AI tool as this may violate the authors' confidentiality and proprietary rights and, where the paper contains personally identifiable information, may breach data privacy rights. This confidentiality requirement extends to the peer review report, as it may contain confidential information about the manuscript and/or the authors. For this reason, reviewers should not upload their peer review report into an AI tool, even if it is just for the purpose of improving language and readability. Peer review is at the heart of the scientific ecosystem and Elsevier abides by the highest standards of integrity in this process. Reviewing a scientific manuscript implies responsibilities that can only be attributed to humans. Generative AI or AI-assisted technologies should not be used by reviewers to assist in the scientific review of a paper as the critical thinking and original assessment needed for peer review is outside of the scope of this technology and there is a risk that the technology will generate incorrect, incomplete or biased conclusions about the manuscript. The reviewer is responsible and accountable for the content of the review report. Elsevier's AI author policy states that authors are allowed to use generative AI and AI-assisted technologies in the writing process before submission, but only to improve the language and readability of their paper and with the appropriate disclosure, as per our instructions in Elsevier's Guide for Authors (opens in new tab/window). Reviewers can find such disclosure at the bottom of the paper in a separate section before the list of references. Please note that Elsevier owns identity protected AI-assisted technologies which conform to the RELX Responsible AI Principles (opens in new tab/window), such as those used during the screening process to conduct completeness and plagiarism checks and identify suitable reviewers. These in-house or licensed technologies respect author confidentiality. Our programs are subject to rigorous evaluation of bias and are compliant with data privacy and data security requirements. Elsevier embraces new AI-driven technologies that support reviewers and editors in the editorial process, and we continue to develop and adopt in-house or licensed technologies that respect authors', reviewers' and editors' confidentiality and data privacy rights. *Generative AI is a type of artificial intelligence technology that can |

| | |
|---|---|
| | produce various types of content including text, imagery, audio and synthetic data. Examples include ChatGPT, NovelAI, Jasper AI, Rytr AI, DALL-E, etc. |
| **Wiley (Publisher)** | Artificial Intelligence (AI), or the use of computers to generate content normally created by humans, is becoming more prevalent in publishing. Use of AI-generated content in a Wiley work will be reviewed on a case-by-case basis, but in general, AI-generated content should only be used in the context of commentary or criticism. If any material contained in your work is created using AI, please inform and identify such AI material to your managing editor (or primary Wiley contact) immediately, including the service used to create the AI material.<br><br>Wiley Peer Review Policy By accepting an invitation to review with a Wiley journal, reviewers agree to act in accordance with generally accepted publication ethics and best practices (including the Ethical Guidelines for Peer Reviewers set forth by the Committee on Publication Ethics COPE). Wiley supports and follows these guidelines as well. The reviewer also understands that good quality manuscripts that were not found to be suitable for a particular journal may be referred to journals in a similar subject area within Wiley's network. Such manuscripts and their peer review reports will be transferred to the receiving journal to expedite any further evaluation and the editor's decision. The reviewer grants Wiley the right to re-use the peer review reports in order to provide publishing services, such as the transfer of manuscripts. This will be done in accordance with both journal and COPE guidelines. The reviewer also consents to the possible transfer of their name, email, and review to a relevant alternate journal. In cases where a manuscript has been transferred from a journal that does not participate in Transparent Peer Review to a journal that does, any reviews submitted to the original journal will not be transferred. Wiley is a member of COPE and is committed to supporting reviewers, authors and editors in ensuring integrity across all aspects of the publishing process. COPE's Core Practicies are at the center of our publication workflows and inform our work in protecting confidentiality throughout peer review. Accordingly, we expect all peer reviewers to respect the confidentiality of peer review and not reveal any details of a manuscript or communications related to it, during or after the peer review process, beyond those that are released by the journal. For journals with single-, double- or triple-anonymized review models, this confidentiality obligation extends to the review and all communications regarding the review. We also require that authors respect the confidentiality of the peer review process, unless the journal has adopted an open review policy. Journals with open review policies will have this noted within their Author Guidelines. |
| **Tylor & Francis (Publisher)** | Taylor & Francis Clarifies the Responsible use of AI Tools in Academic Content Creation The use of artificial intelligence (AI) tools in research and writing is an evolving practice. AI-based tools and technologies include but are not limited to large language models (LLMs), generative AI, and chatbots (for example, ChatGPT). Below we restate our guidance on author accountability and responsibilities as it relates to the use of AI tools in content creation. This policy will be iterated as appropriate. Taylor & Francis recognizes the increased use of AI tools in academic research. As the world's leading publisher of human-centered science, we consider that such tools, where used appropriately and responsibly, have the potential to augment research outputs and thus foster progress through knowledge. Authors are accountable for the originality, validity and integrity of the content of their submissions. In choosing to use AI tools, authors are expected to do so responsibly and in accordance with our editorial policies on authorship and principles of publishing ethics. Authorship requires taking accountability for content, consenting to publication via an author publishing agreement, giving contractual assurances about the integrity of the work, among other principles. These are uniquely human responsibilities that cannot be undertaken by AI tools. Therefore, AI tools must not be listed as an author. Authors must, however, acknowledge all sources and contributors included in their work. Where AI tools are used, such use must be acknowledged and documented appropriately. |

| | |
|---|---|
| **IOP Publishing (Publisher)** | Generative AI (including ChatGPT) IOP Publishing does not accept or condone the use of generative AI, including large language models and AI chatbots such as ChatGPT, to write peer review reports, either fully or partially. By accepting a review invitation, a reviewer agrees to adhere to the ethical standards of IOP Publishing, including reporting any conflicts of interest, ensuring the manuscript under review remains confidential, and retaining their anonymity as a reviewer. Generative AI models are not subject experts as they lack the ability or comprehension to assume responsibility for work they have helped create and are therefore unable to adhere to the ethical standards set out by IOP Publishing. Furthermore, generative AI models do not have the legal personality to sign publishing agreements or licences. Please note that uploading any part of a submitted manuscript to a generative AI model may breach the authors' rights to confidentiality. If a manuscript contains personally identifiable information, it may also breach data protection rights. |
| **American Psychological Association (APA) Publishing (Publisher)** | Generative artificial intelligence, specifically the kind based on Large Language Models (LLMs) like ChatGPT, has become a transformative force in many fields. Scholarly writing and publishing are no different, and generative AI has begun to have an impact on scholarly work. In response to this impact, the APA Publications and Communications Board has approved policies regarding the use of generative AI in scholarly materials. These policies (as well as APA policies on other potential issues in scholarly publishing, and additional reading on the subject) can be found on the APA Publishing Policies page and will continue to develop as we gain a better understanding of the effects of generative AI on scholarly publishing. APA's current policies on generative AI are: When a generative AI model is used in the drafting of a manuscript for an APA publication, the use of AI must be disclosed in the methods section and cited.AI cannot be named as an author on an APA scholarly publication. When AI is cited in an APA scholarly publication, the author must employ the software citation template, which includes specifying in the methods section how, when, and to what extent AI was used. Authors in APA publications are required to upload the full output of the AI as supplemental material. Authors APA policy states that authors are responsible for the accuracy of any information in their article. This means that authors must verify any information and citations provided to them by an AI tool. Authors may use, but must disclose, AI tools for specific purposes such as editing. Please note that for the purposes of this policy, generative AI does not include grammar-checking tools, citation software, or plagiarism detectors which don't employ the use of generative AI; use of these tools does not need to be disclosed or cited in manuscripts submitted to journals. Additionally, please note that when information is entered into generative AI, the organization which runs the generative AI will likely have access to this data. Authors should be aware of this possibility and how it may impact the privacy of participants in their studies, as well as how it may impact their own privacy and intellectual property.<br><br>For this reason, journal editors and reviewers may not enter materials from submitted manuscripts into generative AI as it would constitute a violation of the confidentiality of the peer review process. |
| **IOS Press (Publisher)** | IOS Press follows COPE in stating that AI tools cannot be listed as an author of a paper. The use of AI tools in any step of the research or its reporting must be disclosed in the Materials and Methods (or similar section) of the paper. Authors are fully responsible for the content of their manuscript, even those parts produced by an AI tool, and are thus liable for any breach of publication ethics. |

**Supplementary Table 5. Linear regression analysis of the relationship between journal characteristics and the specificity score of GAI usage guidelines**

| | | Top journals | | | Whole-spectrum sample journals | | |
|---|---|---|---|---|---|---|---|
| | **Journal characteristics** | **Coefficient** | **95% CI** | **P value** | **Coefficient** | **95% CI** | **P value** |
| | | **Specificity score of author and reviewer guidelines** | | | | | |
| **Model 1** | SJR score | -0.03 | (-0.11, 0.05) | 0.49 | 1.21 | (0.72, 1.70) | < 0.01 |
| **Model 2** | Northern America | Ref | - | - | Ref | - | - |
| | Western Europe | -0.28 | (-1.75, 1.19) | 0.70 | 0.05 | (-1.09, 1.18) | 0.94 |
| | Other regions | -4.66 | (-8.91, -0.41) | < 0.05 | -1.39 | (-2.51, -0.26) | < 0.05 |
| | | **Specificity score of author guidelines*** | | | | | |
| **Model 1** | SJR score | -0.03 | (-0.10, 0.05) | 0.47 | 1.21 | (0.72, 1.70) | < 0.01 |
| **Model 2** | Northern America | Ref | - | - | Ref | - | - |
| | Western Europe | -0.29 | (-1.67, 1.09) | 0.68 | 0.05 | (-1.09, 1.18) | 0.94 |
| | Other regions | -4.40 | (-8.39, -0.41) | < 0.05 | -1.39 | (-2.51, -0.26) | < 0.05 |
| | | **Specificity score of reviewer guidelines** | | | | | |
| **Model 1** | SJR score | 0.00 | (-0.02, 0.02) | 0.95 | - | - | - |
| **Model 2** | Northern America | Ref | - | - | - | - | - |
| | Western Europe | 0.01 | (-0.30, 0.31) | 0.97 | - | - | - |
| | Other regions | -0.26 | (-1.14, 0.63) | 0.56 | - | - | - |

Footnote: *For whole-spectrum sample journals, results of "Specificity score of author and reviewer guidelines" are identical to "Specificity score of author guidelines" because specificity score of reviewer guidelines were all zero.

**Supplementary Figure 1. Specificity level of author and reviewer guidelines and requirements of different GAI usage guidelines among whole-spectrum sample journals**

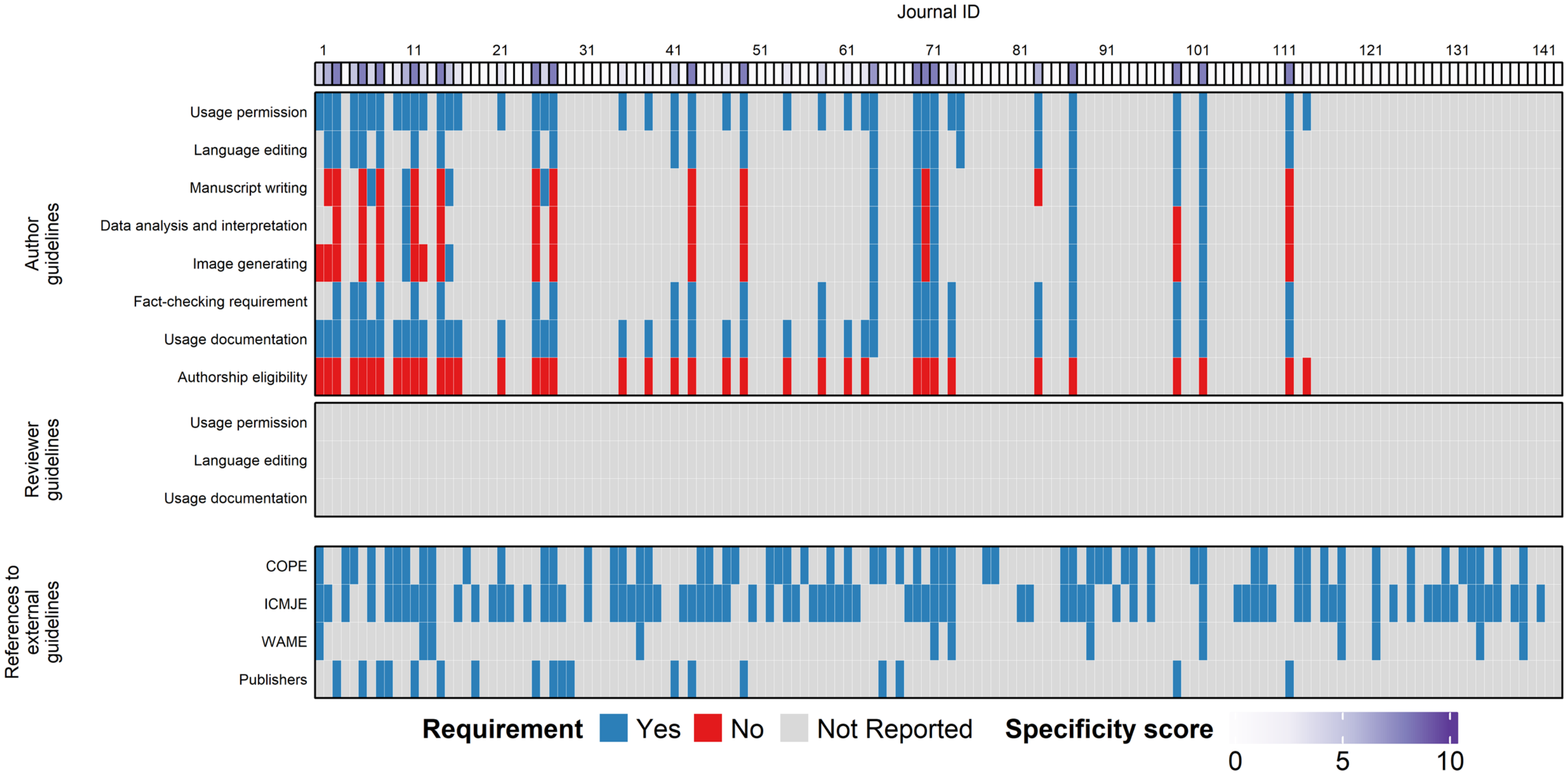

Footnote: The specificity score displays how many requirements were provided for author and reviewer guidelines combined (range: 0-11).
